# Identifying aging and Alzheimer’s disease associated somatic mutations in excitatory neurons from the human frontal cortex using whole genome sequencing and single cell RNA sequencing data

**DOI:** 10.1101/2022.05.25.22275538

**Authors:** Meng Zhang, Gerard A. Bouland, Henne Holstege, Marcel J.T. Reinders

**Affiliations:** Delft Bioinformatics Lab, Delft University of Technology, Delft, The Netherlands; Department of Human Genetics, Amsterdam Neuroscience, Vrije Universiteit Amsterdam, Amsterdam UMC, Amsterdam, The Netherlands; Department of Human Genetics, Leiden University Medical Center, Leiden, The Netherlands

## Abstract

With age, somatic mutations accumulated in human brain cells can lead to various neurological disorders and brain tumors. Since the incidence rate of Alzheimer’s disease (AD) increases exponentially with age, investigating the association between AD and the accumulation of somatic mutation can help understand the etiology of AD. Here we built a somatic mutation detection pipeline by contrasting genotypes derived from WGS data with genotypes derived from scRNA-seq data and applied this pipeline to 76 participants from the ROSMAP project. We focused only on excitatory neurons, the dominant cell type in the human brain. As a result, we identified 196 sites that harbored at least one individual with an excitatory neuron-specific somatic mutation (ENSM) across all individuals, and these 196 sites were mapped to 127 genes. The single base substitution (SBS) pattern of the putative ENSMs was best explained by signature SBS5 from the COSMIC mutational signatures, a clock-like pattern correlating with the age of the individual. The count of ENSMs per individual also showed an increasing trend with age. Among the mutated sites, we found two sites to have significantly more mutations in older individuals (16:6899517 (*RBFOX1*), p = 0.044; 4:21788463 (*KCNIP4*), p = 0.045). Also, two sites were found to have a higher odds ratio to detect a somatic mutation in AD samples (6:73374221 (*KCNQ5*), p = 0.014 and 13:36667102 (*DCLK1*), p = 0.023). 32 genes that harbor somatic mutations unique to AD and the *KCNQ5* and *DCLK1* genes were used for GO-term enrichment analysis. We found the AD-specific ENSMs enriched in the GO-term “vocalization behavior” and “intraspecies interaction between organisms”. Interestingly, we observed both age- and AD-specific ENSMs enriched in the K^+^ channels-associated genes. Taken together this shows our pipeline that combines scRNA-seq and WGS data can successfully detect putative somatic mutations. Moreover, the application of our pipeline to the ROSMAP dataset has provided new insights into the association of AD and aging with brain somatic mutagenesis.

**Author summary:** Somatic mutations are changes in the DNA that occur during life. As with increasing age, somatic mutations also accumulate in human brain cells and can potentially lead to neurological diseases such as Alzheimer’s disease (AD). Associating the occurrence of somatic mutations in human brains with increasing age as well as AD can provide new insights into the mechanisms of aging and the etiology of AD. But somatic mutations do not accumulate similarly across different cell types. Single cell RNA sequencing provides an opportunity to derive somatic mutations for different cell types. We describe a methodology to detect cell-type specific somatic mutations and demonstrate the effectiveness of this methodology by applying it to human brain single cell data of 76 participants from the ROSMAP project. The detected somatic mutational pattern resembles a known clock-like mutational signature, and the number of somatic mutations per person also increases with age. We also identify specific sites that have a higher incidence rate of somatic mutations in AD or associated with increasing age. We further use these findings to postulate molecular pathways enriched with somatic mutations in AD people contributing to the etiology of AD.

## Introduction

Somatic mutations are post-zygotic genetic variations that can result in genetically different cells within a single organism. [1] Possible reasons for the occurrence and accumulation of somatic mutations in human brains are errors occurring during DNA replication and gradual failing of DNA repair mechanisms caused by extensive oxidative stress. [2, 3] Previous studies have shown that brain somatic mutations originating in neuronal stem/progenitor cells can lead to various neurological disorders and brain tumors. [4–6] While mutations in post-mitotic neurons have been found to play an important role in age-related and neurodegenerative diseases, [7] this association remains relatively poorly understood. The link between the accumulation of age-related mutations in neurons and neurodegenerative disease is intuitively worth exploring, considering aging is a major risk factor for many neurodegenerative diseases, like Alzheimer’s disease (AD) [8].

AD is the most predominant form of dementia, characterized by the extracellular accumulation of amyloid beta (Aβ) plaques and the intracellular aggregation of phosphorylated tau protein into neurofibrillary tangles (NFTs). [9, 10] A recent study identified several putative pathogenic brain somatic mutations enriched in genes that are involved in hyperphosphorylation of tau. [11] These results indicate that the aggregation of these neuropathological substrates can be partly explained by the accumulation of brain somatic mutations, which raises a new direction for investigating the pathogenic mechanism of AD.

Most age-related somatic mutations are only present in a small group of post-mitotic neurons or even in a single neuron. For this reason, ultra-deep bulk sequencing and matched peripheral tissues are often required. [11] This type of data is often generated for one specific research question with relatively high cost and are not always available from public databases. In contrast, the availability of public single cell RNA sequencing (scRNA-seq) datasets has exploded due to continuous technological innovations, increasing throughput, and decreasing costs. [12] scRNA-seq data is most often used for expression-based analyses, such as revealing complex and rare cell populations, uncovering regulatory relationships between genes, and tracking the trajectories of distinct cell lineages in development. [13, 14] We hypothesized that scRNA-seq data can also be used to detect somatic mutations.

In this study, we validated the feasibility of calling short somatic variants using scRNA-seq data. We constructed a pipeline to detect brain-specific somatic mutation by contrasting genotypes identified with whole genome sequencing (WGS) data with genotypes identified with scRNA-seq data. For each putative somatic mutation we investigated associated genes and their respective association with AD and age. Additionally, we investigated whether AD and age are associated with an increasing number of somatic mutations.

## Results

### Excitatory neuron-specific somatic mutations (ENSMs)

To study somatic mutations acquired over age and between demented (AD) and non-demented (ND) persons, we collected 90 participants from the ROSMAP data for which WGS data in blood or brain as well as single nuclei RNA sequencing (snRNA-seq) data of the frontal cortex was present (Methods). Since the snRNA-seq data (n=90) were collected from three studies, the read coverage for samples varied between the studies (S1 Fig). To reduce the bias generated from the unbalanced read coverage, we excluded individuals (n=9) with a total read count smaller than 6 × 10^7^. Cells from the snRNA-seq data were annotated according to seven major cell types (Methods). As the amount of cells varied for different cell types (S2 Fig), we first explored the feasibility of detecting somatic mutations for each cell type. This exploratory analysis showed that somatic mutations were only detected from the excitatory neurons (when requiring a minimum number of reads (≥ 5) per sample for a putative variant site, Methods), the dominate cell type in snRNA-seq data. This underpins that a sufficient amount of cells was needed for snRNA-seq based somatic mutation detection. As a consequence, we focus our analysis on excitatory neurons. To further ensure the data quality, we excluded individuals (n=5) which had less than 200 excitatory neurons from our study. After filtering, 76 participants (23 from snRNAseqMFC study, 30 from snRNAseqPFC BA10 study, and 23 from snRNAseqAD TREM2 study) had an adequate read coverage and sufficient number of excitatory neurons. The demographic data (sex, age at death, and cognitive diagnosis (cogdx) categories [15]) of these participants are given in Table 1. More than 72% of them were 85 years of age or older at death; 56% were women. Individuals were grouped based on their cognitive diagnosis in either being non-demented (n=42) or being an AD sample (n=33).

**Table 1.**
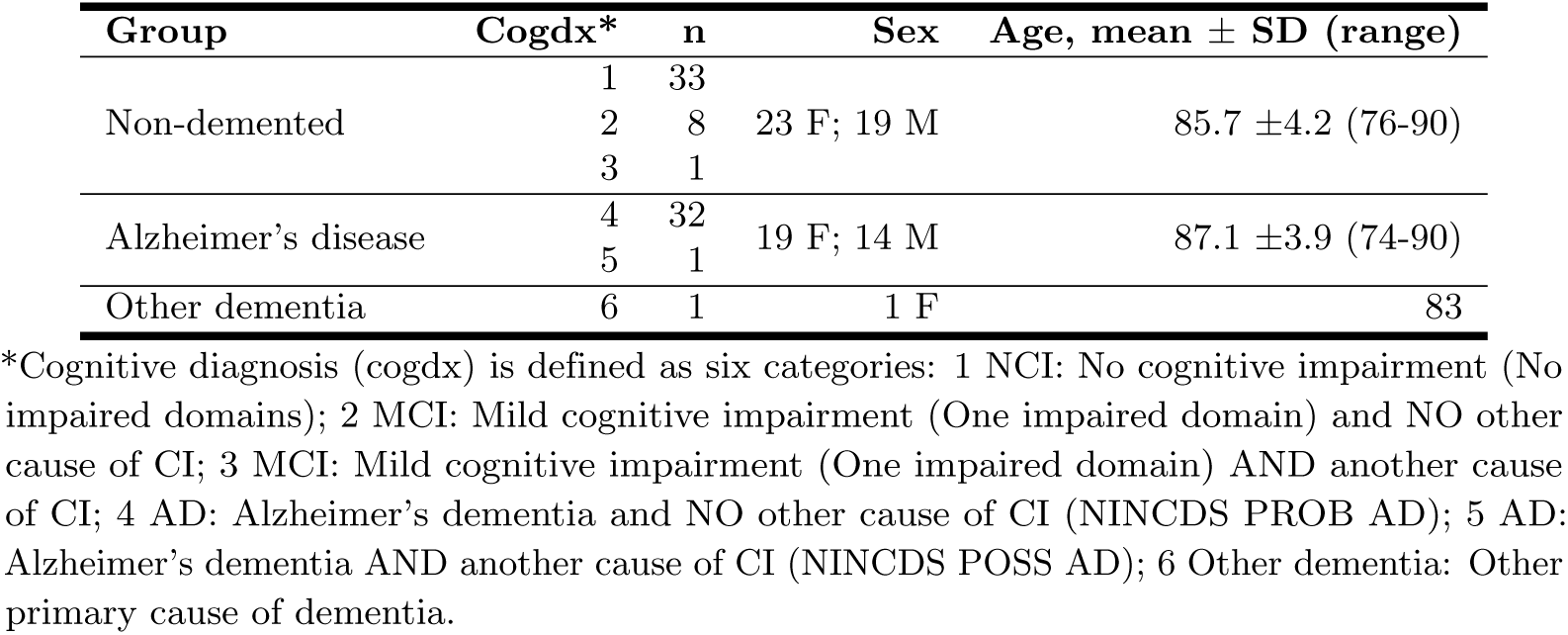
Summary characteristics of selected sample from the ROSMAP study.

| Group | Cogdx* | n | Sex | Age, mean ± SD (range) |
| --- | --- | --- | --- | --- |
| Non-demented | 1 | 33 | 23 F; 19 M | 85.7 ± 4.2 (76-90) |
|  | 2 | 8 |  |  |
|  | 3 | 1 |  |  |
| Alzheimer's disease | 4 | 32 | 19 F; 14 M | 87.1 ± 3.9 (74-90) |
|  | 5 | 1 |  |  |
| Other dementia | 6 | 1 | 1 F | 83 |

\*Cognitive diagnosis (cogdx) is defined as six categories: 1 NCI: No cognitive impairment (No impaired domains); 2 MCI: Mild cognitive impairment (One impaired domain) and NO other cause of CI; 3 MCI: Mild cognitive impairment (One impaired domain) AND another cause of CI; 4 AD: Alzheimer's dementia and NO other cause of CI (NINCDS PROB AD); 5 AD: Alzheimer's dementia AND another cause of CI (NINCDS POSS AD); 6 Other dementia: Other primary cause of dementia.

### Number of detected ENSMs as expected

Somatic mutations in the 76 participants were detected using the pipeline described in the Methods and shown in Fig 1. For that the snRNA-seq data of the excitatory neurons are compared to WGS data of blood (n=23) or brain (n=53). IBD estimation using shared variation sites confirmed the matching between the snRNA-seq and WGS samples (pair-wised PI HAT *>* 0.85, S3 Fig, Methods). From the 9,751,193 short variants called from the snRNA-seq data, we identified 196 sites that harbored at least one sample with an excitatory neuron-specific somatic mutation (ENSM) across all samples. These genetic sites map to 127 genes (Methods), and 104 sites among them were single-nucleotide variants (SNVs). From these 196 sites, 98 were shared between multiple individuals (*n >* 2) (S4 Fig). A few sites have mutations present in almost all individual genomes, which are likely to be either RNA editing events [16]; transcription errors, which can occur in a wide variety of genetic contexts with several different patterns [17, 18]; or technical errors [19]. 53 sites have mutations uniquely present in the brains of the AD samples (S1 Table).

**Fig 1.**
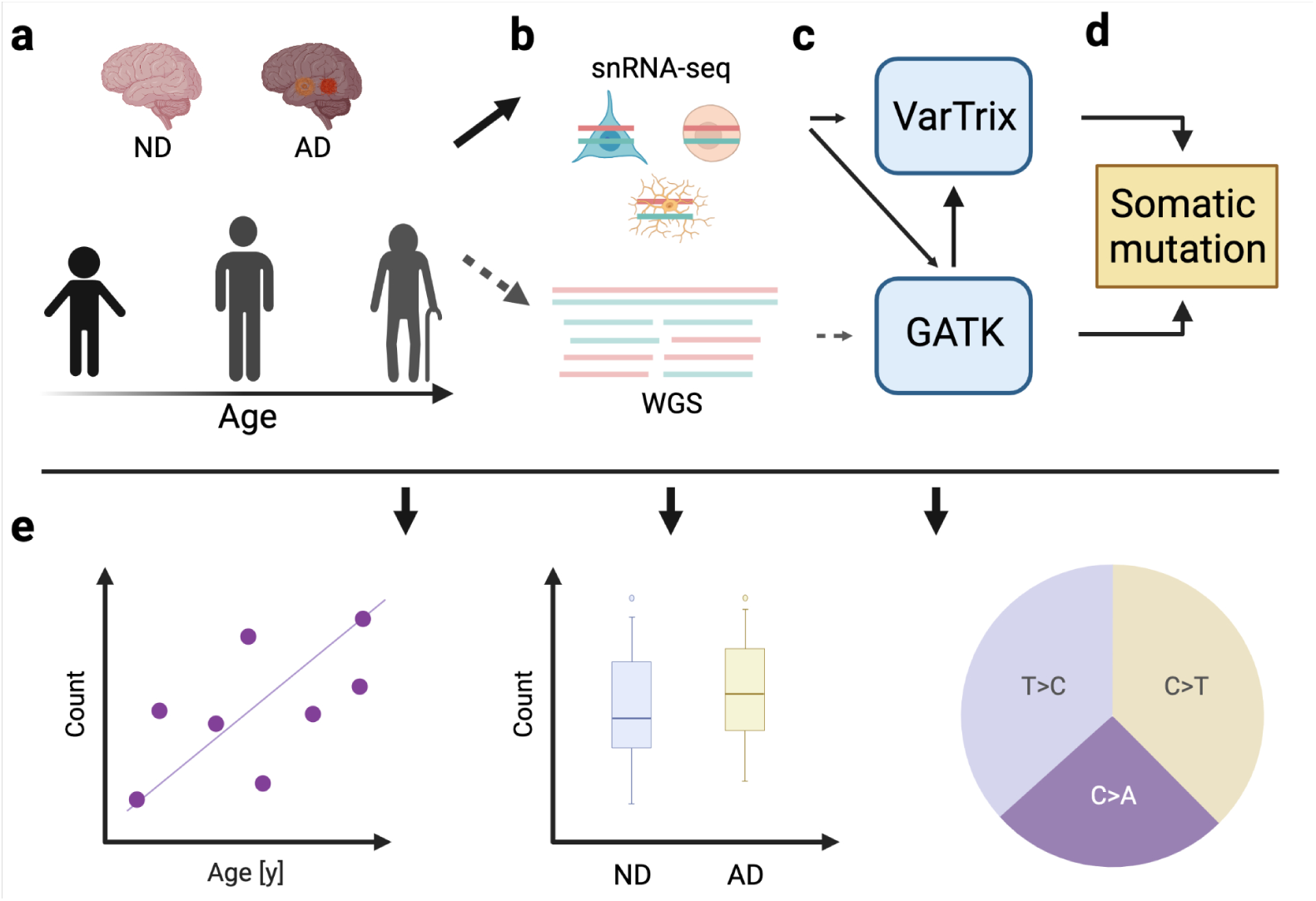
Schematic overview of the analyses in this study. **a**. Participants from the ROSMAP project with both single nuclei RNA sequencing (snRNA-seq) data and whole genome sequencing (WGS) data available were selected for this study. 76 participants who passed the total read number and cell number filtering were used for the analyses. These individuals (aged 74-90) were grouped as non-demented (ND) samples (n=42) or Alzheimer’s disease (AD) patients (n=33) based on the cognitive diagnosis. **b**. The snRNA-seq data were collected from three studies (23 from snRNAseqMFC study, 30 from snRNAseqPFC BA10 study, and 23 from snRNAseqAD TREM2 study). All specimens for these three snRNA-seq data sources were collected post-mortem from the frontal cortex. The snRNA-seq data were clustered and assigned to seven major cell types, but only the reads from excitatory neurons were used for the study after an exploratory phase. The WGS data were measured from blood (n=23) or brain (n=53). **c**. The genotypes for snRNA-seq data were detected using the GATK pipeline followed by VarTrix, while the genotypes for WGS data were detected by the GATK pipeline (Method). **d**. The somatic mutations were determined by contrasting genotypes derived from WGS data with genotypes derived from snRNA-seq data. **e**. For the detected putative somatic mutations, their mutational signature, associated genes, and respective association with AD and age were investigated.

Per individual genome the ENSMs ranged from 24 to 41. This seems to be in agreement with other studies that found an average of 12 somatic SNVs in hippocampal formation tissue using bulk exome sequencing [11], and an average amount of 1700 somatic mutations (substitutions 1500; indels 200) in neurons using a whole-genome duplex single-cell sequencing protocol [20].

### Number of ENSMs increase with age

To characterize the ENSMs, a mutation signature analysis was performed on 104 putative somatic SNVs (Methods). The results show that, from the COSMIC mutational signatures, SBS5 best explains the observed pattern of putative somatic SNVs by Mutalisk (Fig 2). SBS5 is a clock-like signature, i.e. the number of mutations correlates with the age of the individual. This suggests that the underlying mutational processes of the found ENSMs might be part of the normal aging process in excitatory neurons. A previous study using bulk exome sequencing also found an abundance of the SBS5 signature in aged brain tissues. [11]

**Fig 2.**
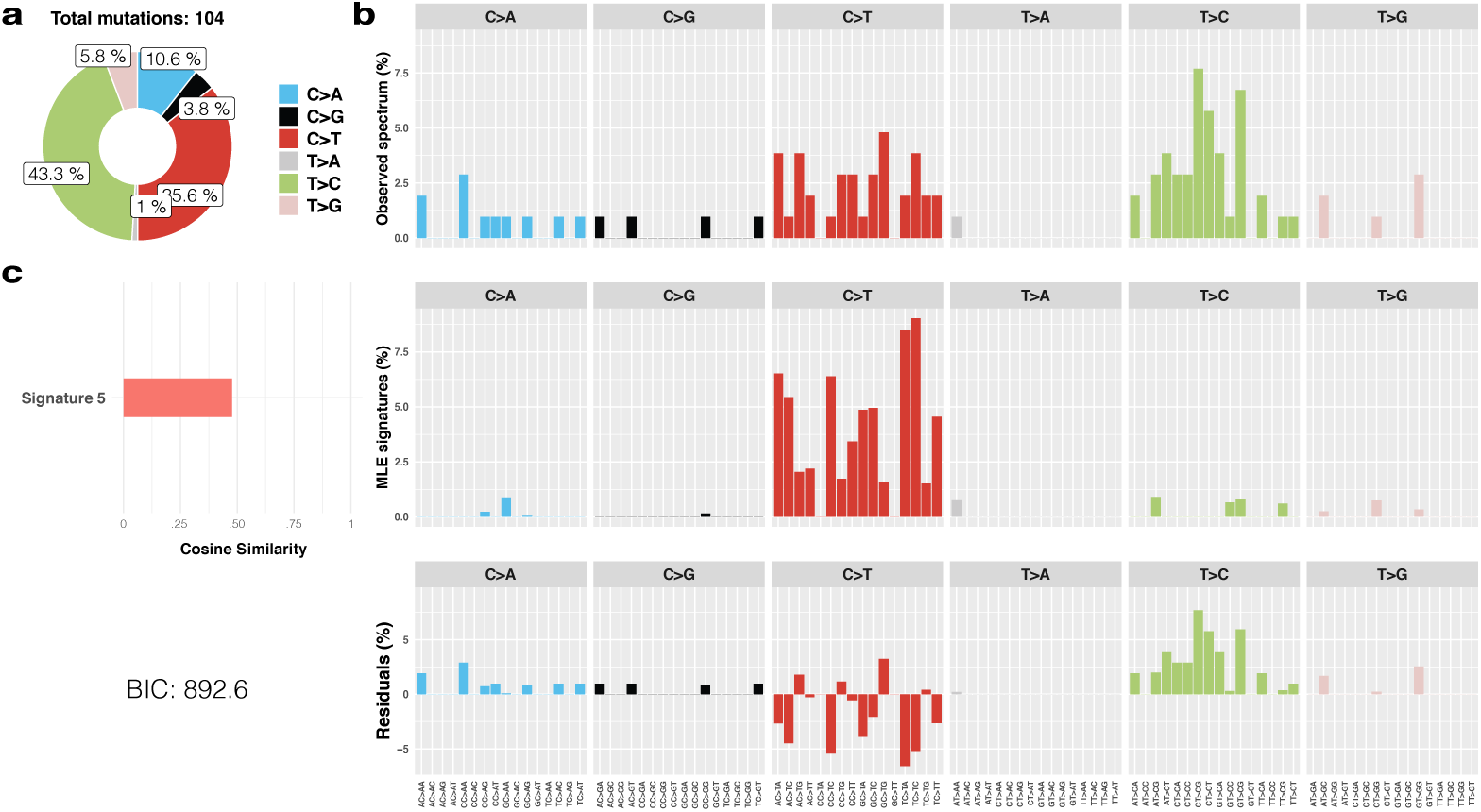
The mutation signature of 104 putative excitatory neuron-specific single nucleotide variations (SNVs) in the brain. Among the 30 COSMIC single base substitution (SBS) signatures, SBS5 was identified as the model that best explains the observed pattern of putative somatic SNVs by Mutalisk (BIC=892.6). **a**. The percentage of each substitution subtype in the 104 putative excitatory neuron-specific SNVs. Subtype T*>*C and C*>*T are the dominate subtypes, account for 43.3% and 35.6% of the fraction separately. **b**. The top panel shows the observed distribution of mutations across the 96 possible mutation types; the middle panel shows the summation of the distributions of the decomposed signatures; the bottom panel shows the difference of each base substitution subtype between the top and middle panel. **c**. The cosine similarity between the distributions of observed mutations and the best decomposition model (SBS 5).

When studying the count of somatic mutation in our analyses, we found only a slight increase with age (*β* = 0.15, Fig 3a) that was not statistically significant (p=0.12). We should note that the number of samples is relatively low and represent a relatively narrow age range (from 74 to 90 years old). Moreover, participants with an age older than 90 years were all censored by age 90, which could also influence the significance of the age trend. A significant trend is observed when we exclude individuals at age 90 from the regression (*β* = 0.37, *P* = 0.005; S5 Fig).

**Fig 3.**
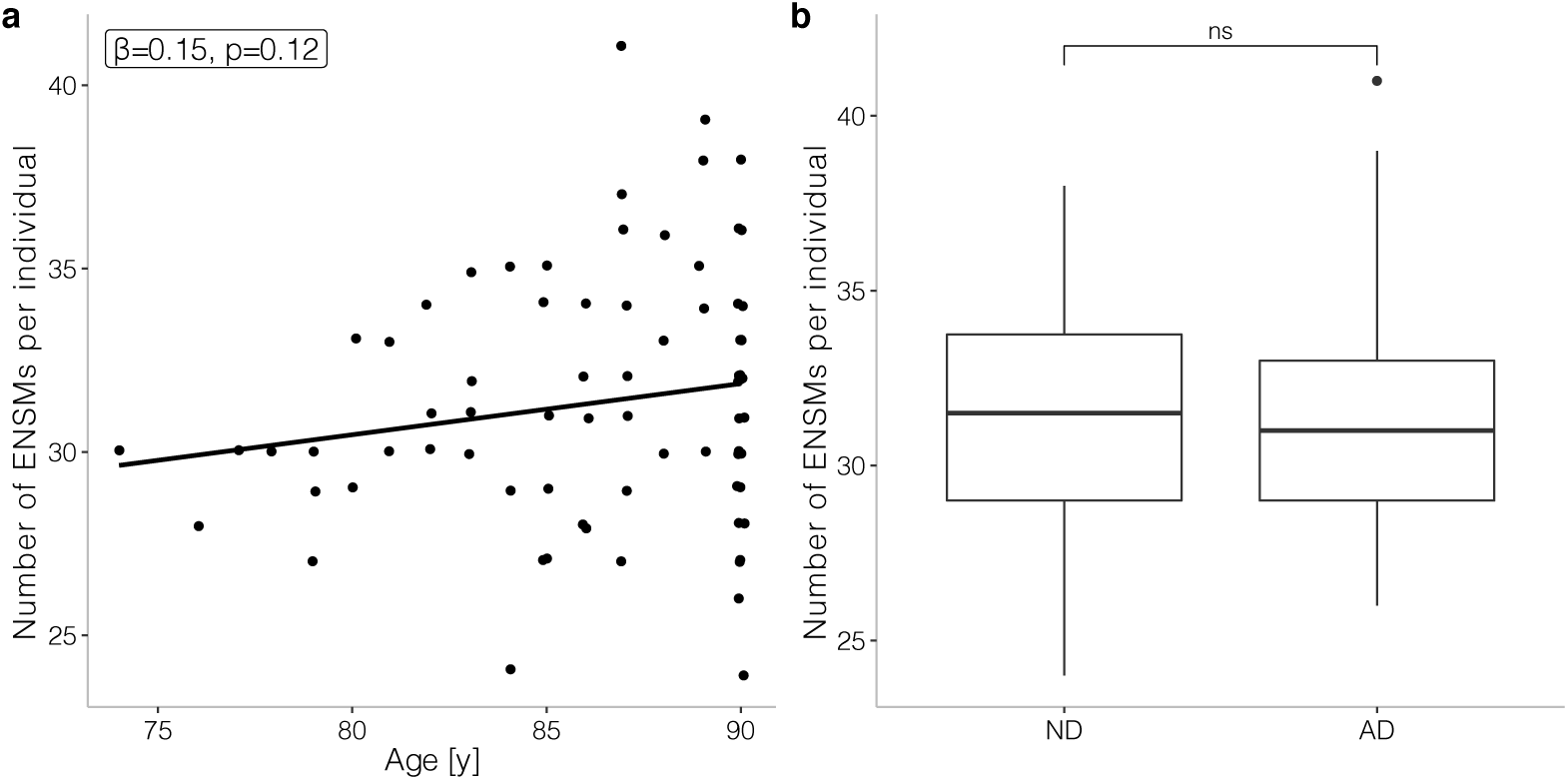
Quantitative comparison of the number of excitatory neuron-specific somatic mutations (ENSMs) in terms of AD and aging. **a**. The number of ENSMs per individual against the age of the individual. The line shows how this number regresses with age. The significance of the coefficient (*β ≠* 0) was tested using t-test. **b**. Boxplot of the number of ENSMs in non-demented controls (ND) and AD patients (AD). The Wilcoxon rank sum test does not show a significance difference (ns).

### *RBFOX1* and *KCNIP4* harbor age-associating ENSMs

As several detected ENSMs are being detected in multiple individual genomes (S4 Fig), we next tested the association of age with somatic mutation prevalence for each site *individually* using a logistic regression (Methods). We added AD status as an explanatory term and excluded the sample with other primary cause of dementia (Methods) from this analysis. Two sites (16:6899517 (*RBFOX1*), *p* = 0.044; 4:21788463 (*KCNIP4*), *p* = 0.045) are found to have significantly more mutations in older individuals. The age distributions in mutated and unmutated samples for these two sites are shown in Fig 4. Some caution should be treated when interpreting this plot for individuals older than 90 years as these are all mapped to 90 years old.

**Fig 4.**
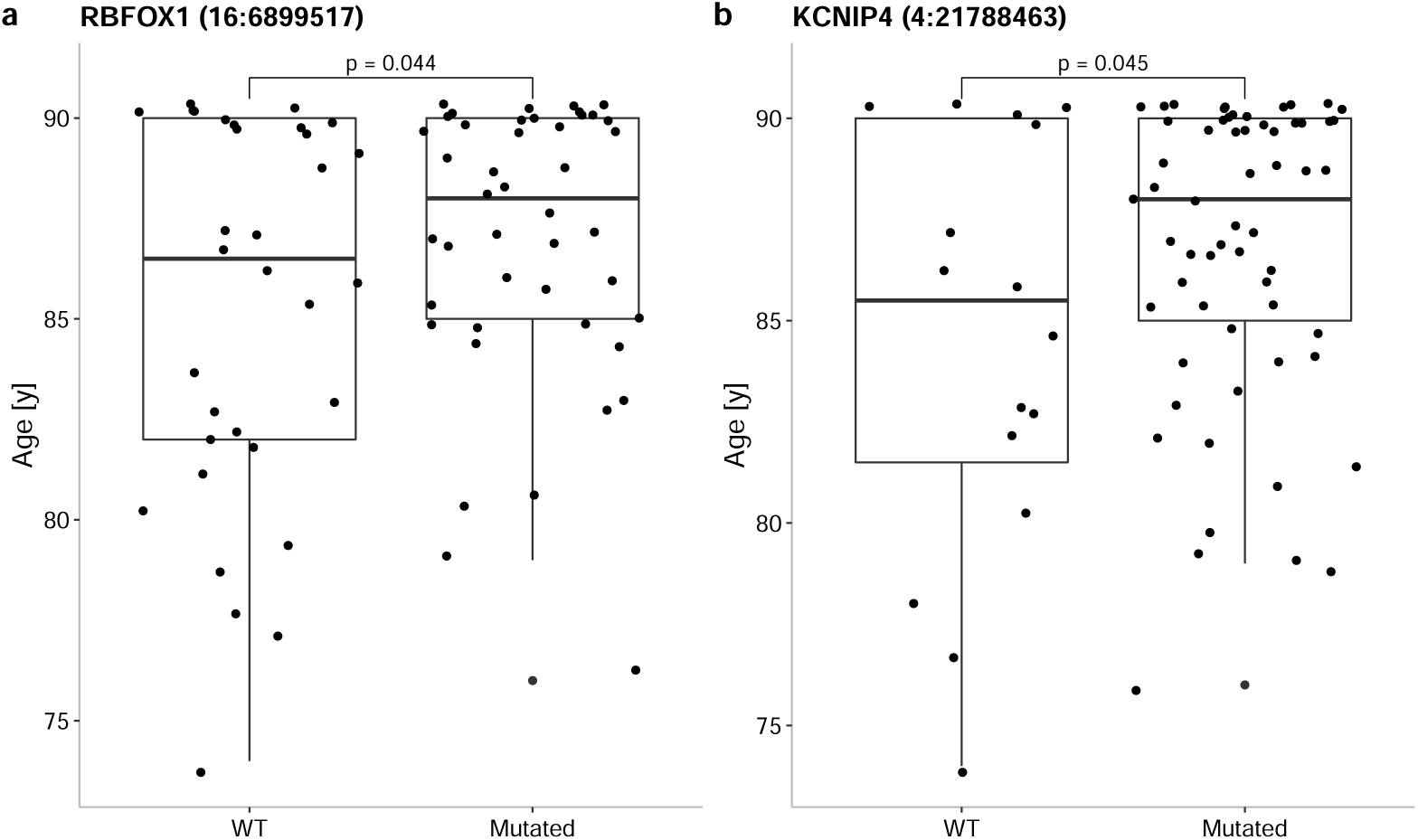
The occurrence of somatic mutation with age in (a) *RBFOX1* and (b) *KCNIP4* genes. Logistic regression was used to test the prevalence of somatic mutations with increasing age.

### ENSM sites in *KCNQ5* and *DCLK1* associate with AD status

Genes that were enriched with somatic mutations in AD samples might have a higher possibility to be associated with AD. We found 53 ENSM sites that were only detected in AD samples. This prompted the question whether the number of ENSMs associate with AD status. A Wilcoxon rank sum test indicated that there was no significant difference (p=0.71) in the average count of ENSMs between AD samples and non-demented controls (Fig 3b). This finding is in line with a previous report [11, 21] that indicated that somatic mutations are associated with AD in certain patterns, but not by amount.

Next, we examined whether the occurrence of an ENSM is overrepresented within AD samples. A Fisher’s exact test that identifies sites that have a higher odds ratio to detect a somatic mutation in AD samples (Methods), yielded two sites with significant odds ratios. These sites are mapped to two genes (6:73374221 (*KCNQ5*), *p* = 0.014 and 13:36667102 (*DCLK1*), *p* = 0.023).

### Genes harboring AD specific ENSMs do relate to Alzheimer or processes involved in Alzheimer

The 53 AD specific ENSM sites map to 42 genes. When we exclude genes for which also an ENSM occurs in an ND individual (n=10), we end up with 32 genes that have ENSMs only seen in AD samples (S1 File). Among these 32 genes, there are several well-known AD-associated genes, like *SLC30A3, TTL*, and *CTSB*, which thus harbor somatic mutations unique for AD.

Together with the two genes for which AD samples had a higher occurrence of ENSMs (*KCNQ5* and *DCLK1*), we conducted a GO-term analysis to investigate the biological pathways that may be involved (Methods). The most enriched biological processes is “vocalization behavior” (*FDR <* 0.009) which is known to be associated with AD. [22] Also, “intraspecies interaction between organisms” is found to be significant (*FDR <* 0.042), and detected genes with this function are *DLG4, CNTNAP2*, and *NRXN3* (Fig 5). Our results also identified a group of genes (*CACNA1B, CNTNAP2, DLG4, KCNQ3*, and *KCNQ5*) enriched with the GO-term “ion channel complex” (*FDR <* 0.031). *KCNQ* genes encode five members of the K_v_7 family of K^+^ channel subunits (K_v_7.1–7.5). Four of these (K_v_7.2–7.5) are expressed in the nervous system. [23] Concerning AD-related neuropathology, a link between Aβ accumulation and K_v_7 channels has been reported by some studies. [24, 25]

**Fig 5.**
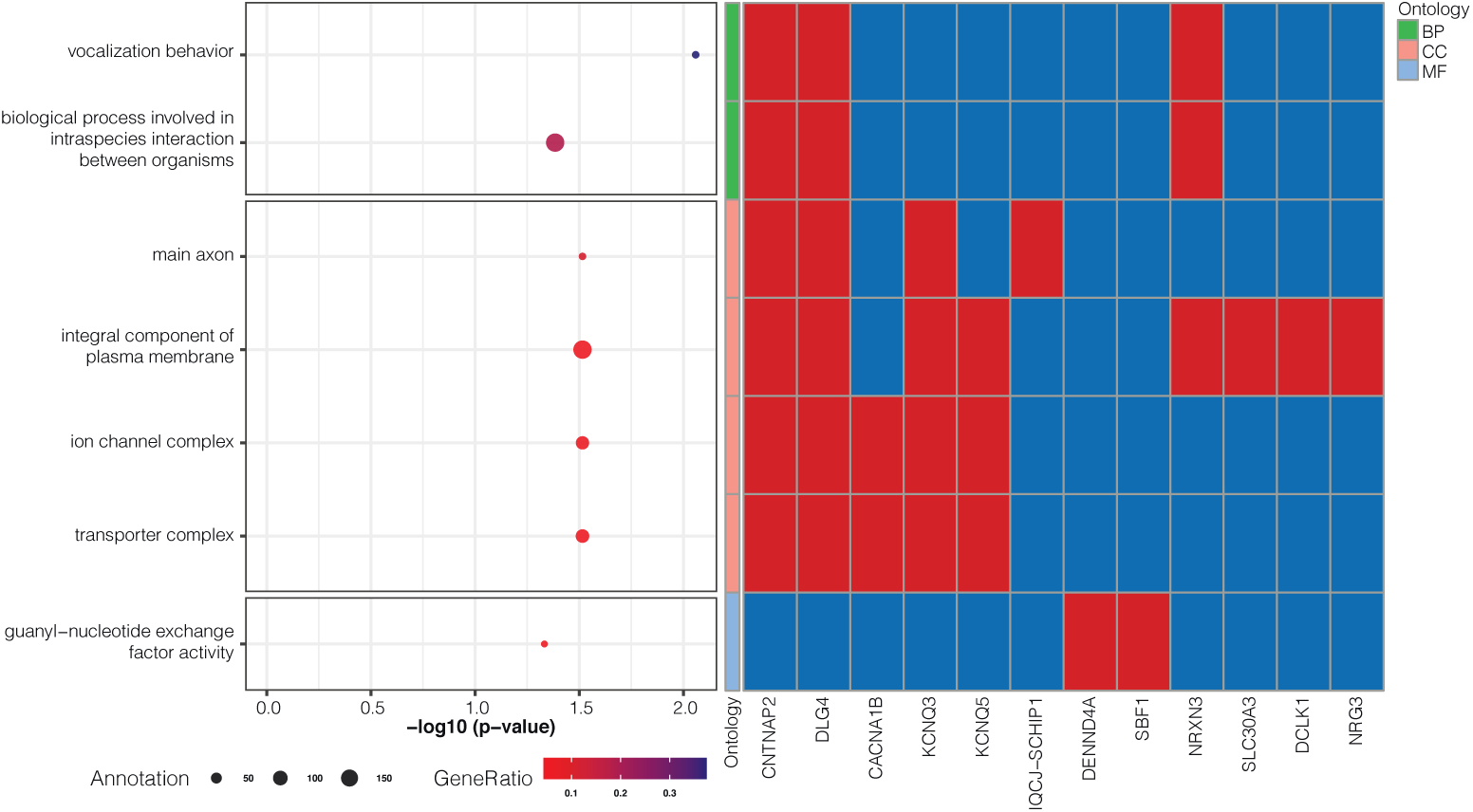
GO-terms enriched with genes having AD-specific ENSMs. 32 genes that have ENSMs only seen in AD samples, and the *KCNQ5* and *DCLK1* genes that have a higher occurrence in AD samples are used in the GO-term enrichment analysis. The left panel of the figure shows the enriched terms, their corrected p-value, the number of genes annotated with that term (size of circle), and the fraction of overlapping genes that harbor a AD-specific ENSM (color of circle). The FDR corrected significant GO-terms are grouped into three categories: Biological Process (BP), Cellular Component (CC), and Molecular Function (MF). The right panel shows the subset of genes having an AD-specific ENSM that are annotated with the enriched GO terms, red squares, while a blue squares indicates that the gene does not have that annotation. Those genes that are not annotated with any of these GO-terms are not included in this panel.

## Discussion

Late-onset Alzheimer’s disease, whose incidence increases with age, is often referred to as a geriatric disease. Although the accumulation of Aβ peptide and phosphorylated tau protein are the neuropathological main characteristics of AD, they fail to explain the molecular pathogenesis. As such, a cell-level investigation might be necessary to study the underlying pathogenic mechanism. Here, we identified somatic mutations using public data collected from 76 ROSMAP donors and investigated their associations with AD and aging.

Although scRNA-seq data are normally used for expression-based analyses, our results have shown that scRNAseq data can be used for the detection of somatic mutations at a cell-type specific level. As long as RNA sequences align correctly to a reference genome, the pipeline that was used for variant calling can be used for both bulk RNA-seq and scRNAseq data. [26] However, calling variants for each cell separately is not efficient, suffers from low coverage, and each cell is likely to have a unique set of identified variants. For this reason, we aggregated cells per individual and per cell-type, generating cell-type specific pseudo-bulk data. An exploratory run of the pipeline revealed that this approach was only feasible for excitatory neuron as it was the most abundant cell type in the snRNA-seq data resulting in sufficient coverage. This observation shows that a sufficient amount of cells or relatively deep sequencing is required for somatic mutation detection.

Our analysis showed that the prevalence of somatic mutations in the gene *KCNIP4* and *RBFOX1* is associated with increasing age (when corrected for AD status). *KCNIP4* encodes a member of the family of voltage-gated potassium (K^+^) channel-interacting proteins (*KCNIPs*), which suggests altered ion transports/channels may be associated with the aging process. [27] *RBFOX1* is a neuron-specific splicing factor predicted to regulate neuronal splicing networks clinically implicated in neurodevelopmental disorders. [28, 29] The increased somatic mutations in *RBFOX1* with age indicates neurodevelopmental disorders may also associate with human brain aging.

We detected the occurrence of somatic mutations close to well-known AD-associated genes, like *SLC30A3, TTL*, and *CTSB. SLC30A3* is known to be down-regulated in the prefrontal cortex of AD patients. [30] *SLC30A3* is assumed to play a protective role against ER stress, which has been thought to be involved to neurodegenerative diseases such as AD. [31] *TTL* is a cytosolic enzyme involved in the post-translational modification of alpha-tubulin. [32] A previous study found that levels of *TTL* were decreased in lysates from AD brains compared to age-matched controls and that, in contrast, D2 tubulin was significantly higher in the AD brains, indicating that loss of *TTL* and accompanying accumulation of D2 tubulin are hallmarks of both sporadic and familial AD. [33] Gene *CSTB* encodes cystatin B (CSTB), an endogenous inhibitor of cystine proteases. [34] Human CSTB has been proposed to be a partner of Aβ and colocalises with intracellular inclusions of Aβ in cultured cells. [35] Protein levels of CSTB have been also reported to increase in the brains of AD patients. [36] These observations suggest that somatic mutations may initiate or are involved in the AD process in many ways.

Advance AD-related dementia is often accompanied with language problems, behavioral issues and cognitive decline. [8] Our results identified AD associated somatic mutations in the genes *CNTNAP2, DLG4*, and *NRXN3* which are involved in, among other processes, vocalization behavior, intraspecies interaction between organisms and behavior and cognition. Our results show that AD-related language problems and behavioral issues might be associated with somatic mutations in excitatory neurons. In addition, we identified AD-associated somatic mutations in *CACNA1B, CNTNAP2, DLG4, KCNQ3* and *KCNQ5*, which are all ion-channels or involved with ion-channels. Previous studies have reported on the possible role of altered neuronal excitability, controlled by different ion channels and their associated proteins, occurring early during AD pathogenesis. [37, 38] Specifically K^+^ channels which are the most numerous and diverse channels present in the mammalian brain, may partly explain this alteration in neuronal excitability. [39] Also, a dysfunction of K^+^ channels has been observed in fibroblasts [40] and platelets [36] of AD patients. Additionally, Aβ has been demonstrated to not only be involve in the AD pathogenesis, but also modulate K^+^ channel activities [41] and may have a physiological role in controlling neuronal excitability [42]. Somatic mutations involved in K^+^ channels were detected to associate with both AD and age indicating the existence of common processes behind neurodegenerative disease and aging. It also seems that K^+^ channels are naturally subjected to oxidation by reactive oxygen species (ROS) in both aging and neurodegenerative disease which are characterized by high levels of ROS. [43]

Calling variants and detecting somatic mutations from public scRNA-seq data expands the use scope of scRNA-seq data, and may provide new insight into postzygotic genetic change at a cell-type specific level. The use of a single cell-type (excitatory neuron) and the minimal read coverage requirement minimized biases driven by gene-specific expression. However, some limitations can also not be ignored. With the pipeline that was used, the results are sensitive to the chosen settings of the parameters. RNA editing events and transcription errors that happen in RNA sequences might also be identified as somatic mutations using this pipeline, but the association between this type of mutation and AD or aging could also be interesting. [44] Additionally, as the pipeline is relatively complex, quality control was highly critical for this study. Another limitation of this study is the relative narrow age range of the included individuals. Moreover, ages above 90 were censored to be 90. These two factors may explain that we only found a relative weak association between age and the accumulation of somatic mutations. On the other hand, the significant trend after removing individuals with an age higher than 90 might also indicate that nonagenarians and centenarians generally have a more healthy individual genome. Next, heterozygous variants from the WGS data were ignored in this study (due to potential ambiguity due to differences in gene expression). Therefore, many potential somatic mutation were excluded from the start. Finally, as 10x scRNA-seq data was used to detect somatic mutations, only variants located on the DNA that gets transcribed into mRNA were detected.

Our study has explored the feasibility of using scRNA-seq data to detect somatic variants. We developed a pipeline that combines scRNA-seq and WGS data and successfully detected putative somatic mutations, as such, broadened the use-scope of scRNA-seq data. By applying this pipeline on data obtained from the ROSMAP project, we generated potential new insights into the association of AD and aging with brain somatic mutagenesis. It should be noted that follow-up studies with larger cohorts are required to validate our results.

## Materials and methods

### Case selection

Single nuclei RNA sequencing (snRNA-seq) data and whole genome sequencing (WGS) data were obtained from the Religious Order Study (ROS) and the Rush Memory and Aging Project (MAP), two longitudinal cohort studies of aging and dementia. [45] Information collected as part of these studies, collectively known as ROSMAP, includes clinical data, detailed post-mortem pathological evaluations and tissue omics profiling. The snRNA-seq data used in this project were from three sources: 1) snRNAseqMFC study (n=24), 2) snRNAseqAD TREM2 study (n=32) [46], and 3) snRNAseqPFC BA10 study (n=48) [47]. All specimens for these three snRNA-seq data sources were collected post-mortem from the frontal cortex, sub-regions might slightly differ between studies. The snRNA-seq data from the three studies were all sequenced according to the 10x Genomics manufacturer’s protocol. The detailed information for cell partitioning, reverse transcription, library construction, and sequencing run configuration for the three studies is available on Synapse (snRNAseqMFC: syn16780177, snRNAseqAD TREM2: syn21682120, snRNAseqPFC BA10: syn21261143). WGS data was from a subset of the ROSMAP participants with DNA obtained from brain tissue, whole blood or lymphocytes transformed with the EBV virus. The details for WGS library preparation and sequencing, and WGS Germline variants calling were described in the previous study. [48] The individuals (n=90) that have both snRNA-seq data and WGS data (27 from brain tissue and 63 from whole blood) available were selected for this study. Individuals annotated with no cognitive impairment or mild cognitive impairment were defined as non-demented (ND) controls; AD patients with or without other cause of cognitive impairment were defined as AD samples.

### Cell type annotation

Each snRNA-seq dataset was separately processed for clustering and cell type annotation which was done as follows. The processed count matrix was loaded in Seurat 3.2.2. [49] The data was log-normalized and scaled before analysis. Next, with the 2,000 most variable genes (default with Seurat), principal components analysis (PCA) was performed. The number of principal components used for clustering was determined using the elbow method. Further, Seurat’s FindNeighbours and FindCluster functions were used, which utilizes Louvain clustering, the resolution was set at 0.5. A UMAP plot (S6 Fig) was made to visualise and inspect the clusters. The following cell types were identified using known and previously used markers: excitatory neurons (*SLC17A7, CAMK2A, NRGN*), inhibitory neurons (*GAD1, GAD2*), astrocytes (*AQP4, GFAP*), oligodendrocytes (*MBP, MOBP, PLP1*), oligodendrocyte progenitor cell (*PDGFRA, VCAN, CSPG4*), microglia (*CSF1R, CD74, C3*) and endothelial cells (*FLT1, CLDN5*). [47] Based on the markers’ expression patterns across clusters determined by Seurat’s FindMarkers function, cell types were assigned to cells (S2 File). When clusters were characterized by markers of multiple cell types, they were assigned “Unknown”.

### snRNA-seq short variants calling

Single nuclei RNA reads were mapped to the reference human genome GRCh37 using STAR aligner (STAR v2.7.9a). After alignment, duplicate reads were identified using MarkDuplicates (Picard v2.25.0) and reads with unannotated cell barcodes were removed using samtools (smatools v1.11). Reads containing Ns in their cigar string were splitted into multiple supplementary alignments using SplitNCigarReads (GATK v4.2.0.0) to match the conventions of DNA aligner. Base Quality Recalibration was performed per-sample to detect and correct for patterns of systematic errors in the base quality scores using BaseRecalibrator and ApplyBQSR (GATK v4.2.0.0). Short variant discovery was performed on chromosome 1-22 with a two-step process. HaplotypeCaller was run on each sample separately in GVCF mode (GATK v4.2.0.0) producing an intermediate file format called gVCF (for genomic VCF). gVCFs from each individual were combined together and run through a joint genotyping step (GATK v4.2.0.0) to produce a multi-sample VCF file. S7 Fig indicats the steps of snRNA-seq short variants calling in a flow chart. Variant filtration was then performed using bcftools (bcftools v1.11). A basic hard-filtering was performed using cutoffs of 1) the total read depth DP*<*50000; 2) the quality of calling QUAL*>*100; 3) the quality by depth QD*>*2; 4) the strand odds ratio SOR*<*2; 5) the strand bias Fisher’s exact test FS*<*10.

### Identical individual check using IBD estimation

To make sure the sequences of snRNA-seq and WGS are matching and from the same individual, we performed a pairwise identical by descent (IBD) estimation using filtered variants from snRNA-seq and WGS in a combined VCF file. The estimation was calculated using PLINK v1.9. [50] The proportion IBD value PI HAT from the output of PLINK was used as the estimator, when the profiles are from the same individual the PI HAT value will be close to 1, otherwise it will be close to 0.

### Somatic mutation detection using VarTrix

VarTrix, a software tool for extracting single cell variant information from 10x Genomics single cell data, was used to detect somatic mutations. For single nuclei gene expression data, VarTrix requires a pre-called variant set in VCF format, an associated set of alignments in BAM or CRAM format, a genome FASTA file, and a cell barcodes file produced by Cell Ranger as input. After an exploratory phase, we observed that only cells annotated as excitatory neuron had enough read coverage for somatic mutation detection. Therefore, for each individual, a subset of the BAM file including only reads from cells annotated as excitatory neuron was used as the input of VarTrix. Correspondingly, the pre-called variant set was also detected from the subset of the BAM file which only including barcodes from cells annotated as excitatory neuron.

Human reference genome GRCh37 was used as the genome FASTA file. In this study, VarTrix was run in coverage mode generating a reference coverage matrix and an alternate coverage matrix indicating the number of reads that support the reference allele and the alternate allele. These matrices were later used for filtering variant sites and detecting somatic mutations in the excitatory neurons. Since the snRNA-seq data were collected from three studies, the average coverage varied between different sources. To minimize the batch effect from different studies, we filtered the variant site based on the read number of each individual. Specifically, we calculated a cutoff *C*_*i*_ for each individual *i* as below:

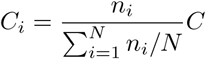

where *n*_*i*_ is the number of reads for individual *i, N* is the number of individuals.

The constant value *C* is set as 25 to guarantee a relatively large minimum average read coverage across individuals. A variant site would be used for somatic mutation detection when for all individuals the read depth at this site is higher than the cut-off *C*_*i*_ for that individual. Next, a somatic mutation was identified as present in one individual when: 1) the genotype of this individual at the site in WGS was ref/ref and the ratio of reads that support the alternate allele is larger than 0.1 at the same site, or 2) the genotype of this individual at the site in WGS was alt/alt and the ratio of reads that support the reference allele is larger than 0.1 at the same site. When the genotype of a individual at a certain site was heterozygote in WGS, we ignored the site for that individual, regardless of the allele ratio in snRNA-seq, because we cannot distinguish an observed homozygous variant at a site in snRNA-seq is due to somatic mutagenesis or reads missing when there is a heterozygous variant in WGS at the same site.

### Mutation signature analysis

To characterize the contribution of mutation signatures, we pooled all putative somatic single nucleotide variations (SNVs) for signature analysis. We formatted the pooled SNVs in a VCF file and used it as input for running Mutalisk [51] with the following configurations: maximum likelihood estimation (MLE) method; linear regression. The input file was compared with 30 single base substitution (SBS) signatures from the COSMIC mutational signatures database. The best model of signature combination was suggested from the tool by considering the Bayesian information criterion (BIC).

### Variants annotation and effect prediction

The gene annotation and functional effect prediction for all putative variants were performed using SnpEff (SnpEff v5.0). [52] The human genome GRCh37 was used as reference genome. If there were multiple genes mapping to one variant site, the gene having higher putative effect was used for the disease and age association analyses.

### GO-term enrichment analysis

The gene ontology (GO-term) enrichment analysis was performed using “topGo” package [53] in R and compressed by REVIGO [54] with semantic similarity score “Lin” [55]. The genes that were annotated to the variant sites with read depths higher than the cut-offs for all samples were used as background. The p-values from the uneliminated GO-terms were corrected using “Benjamini & Hochberg” method, significant results were reported with false discovery rate (FDR) *<* 0.05.

### Statistical analysis

All calculations were performed using R (version 3.6.3). Wilcoxon rank sum test, linear regression, Fisher’s exact test, and logistic regression were performed using the “stats” R package. [56] By categorizing the “present” of somatic mutation as 1 and the “absent” of somatic mutation as 0, the logistic function was defined as: *p* = 1*/*(1 + *exp*(−(*β*_0_ + *β*_1_*age* + *β*_2_*group*))), where *age* is the age of the sample at death, *group* is the assigned group for the individual based on the cogdx category, and *β*_0..2_ are the coefficients of the intercept and the explanatory variables. For this analysis, only individuals from the AD and ND group were used.

## Data Availability

All data produced in the present study are partly available upon reasonable request to the authors.

## Acknowledgments

We thank and acknowledge all participants and their family members of the ROSMAP study. We thank everyone in our life who has supported us both materially and spiritually.

## Author Contributions

MZ, GAB, and MJTR conceived and designed the experiments. MZ and GAB performed the experiments and analyzed the data. MZ, GAB, HH, and MJTR wrote the manuscript. MJTR supervised the research. All authors read and approved the manuscript.

## Supporting information

**S1 Fig. The distribution of the number of single-nuclei RNA (snRNA) reads across individuals**. The dashed red line indicates the cutoff of *<* 6 × 10^7^, individuals below this line were excluded from the study. The colors indicated the study that the individual was from. Individuals with color blue and red were from the two batches (B1 and B2) of snRNAseqMFC study; individuals with color orange were from snRNAseqAD BA10 study; individuals with color purple were from snRNAseqPFC TREM2 study.

**S2 Fig. The number of cells per cell type per individual**. The cell types were distinguished with seven different colors. Individuals with color blue and red were from the two batches (B1 and B2) of snRNAseqMFC study; individuals with color orange were from snRNAseqAD TREM2 study; individuals with color purple were from snRNAseqPFC BA10 study.

**S3 Fig. IBD estimation between paired genetic profiles from WGS and snRNA-seq**. The IBD estimation was performed to ensure the WGS and snRNA-seq profiles which share the same identifier were from the same individual. PI HAT is a measure of overall IBD alleles. If the genetic profiles are from different persons, the value PI HAT will be close to 0. On the contrary, if the profiles are from the same person, the value PI HAT will be close to 1.

**S4 Fig. Distribution of sample counts with excitatory neuron-specific somatic mutations at the same site**.

**S5 Fig. The count of mutations regressed with age**. Individuals at age 90 were removed from this figure. The significance of the coefficient (*β* ≠ 0) was tested using t-test.

**S6 Fig. The UMAP plot for cell type clustering in each study**. See Method for details.

**S7 Fig. The pipeline of short snRNA-seq variants calling**. This pipeline follows the best practices workflows from GATK.

**S1 File. Supplementary file for AD specific ENSM sites**.

**S2 File. Supplementary file for cell type annotation**.

**S1 Table. Counting of mutagenic sites based on the cognitive diagnosis**.

