## Supplementary figures and images for "Identifying aging and Alzheimer’s disease associated somatic mutations in excitatory neurons from the human frontal cortex using whole genome sequencing and single cell RNA sequencing data"

### S1 Fig

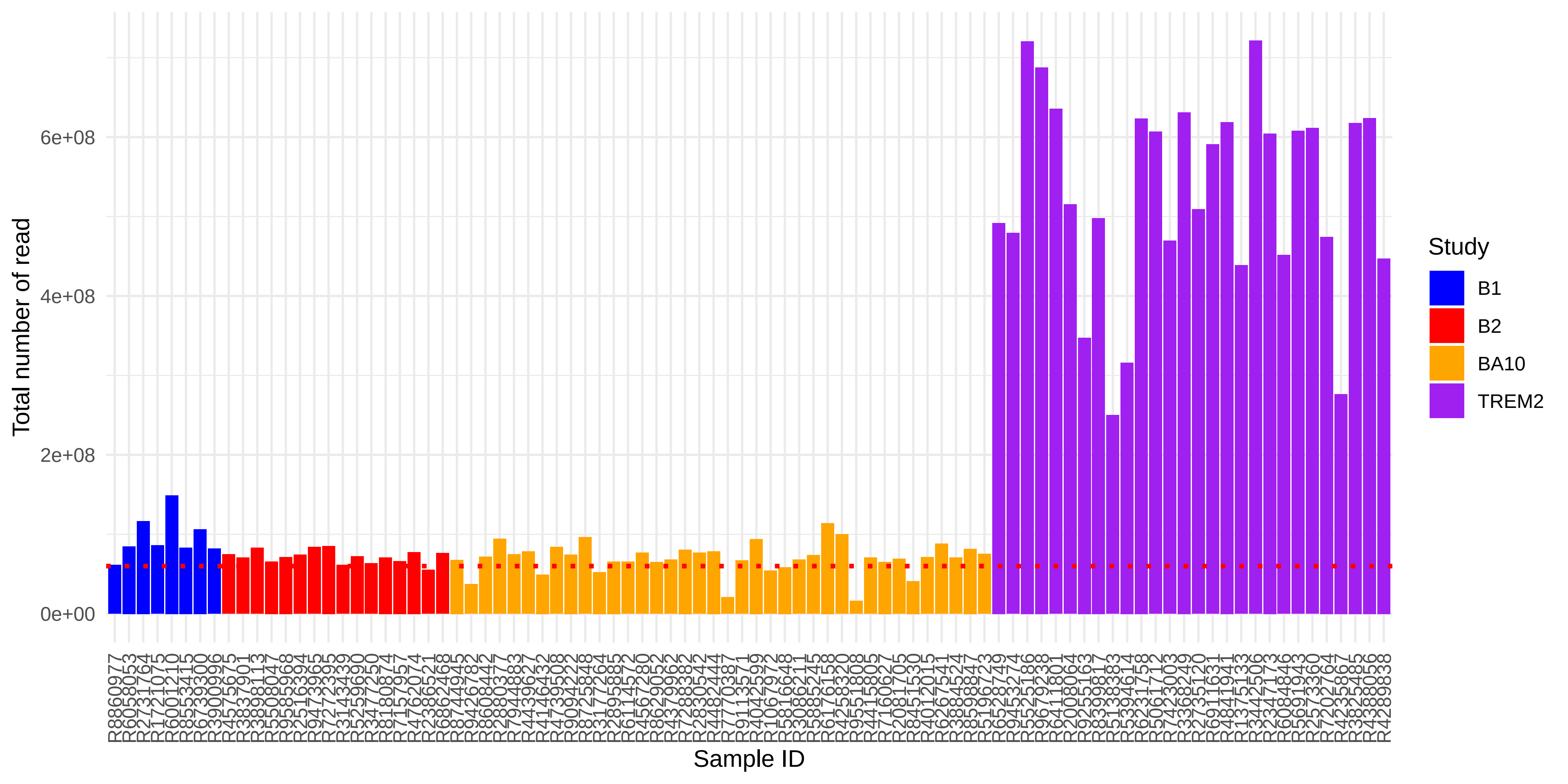

### S2 Fig

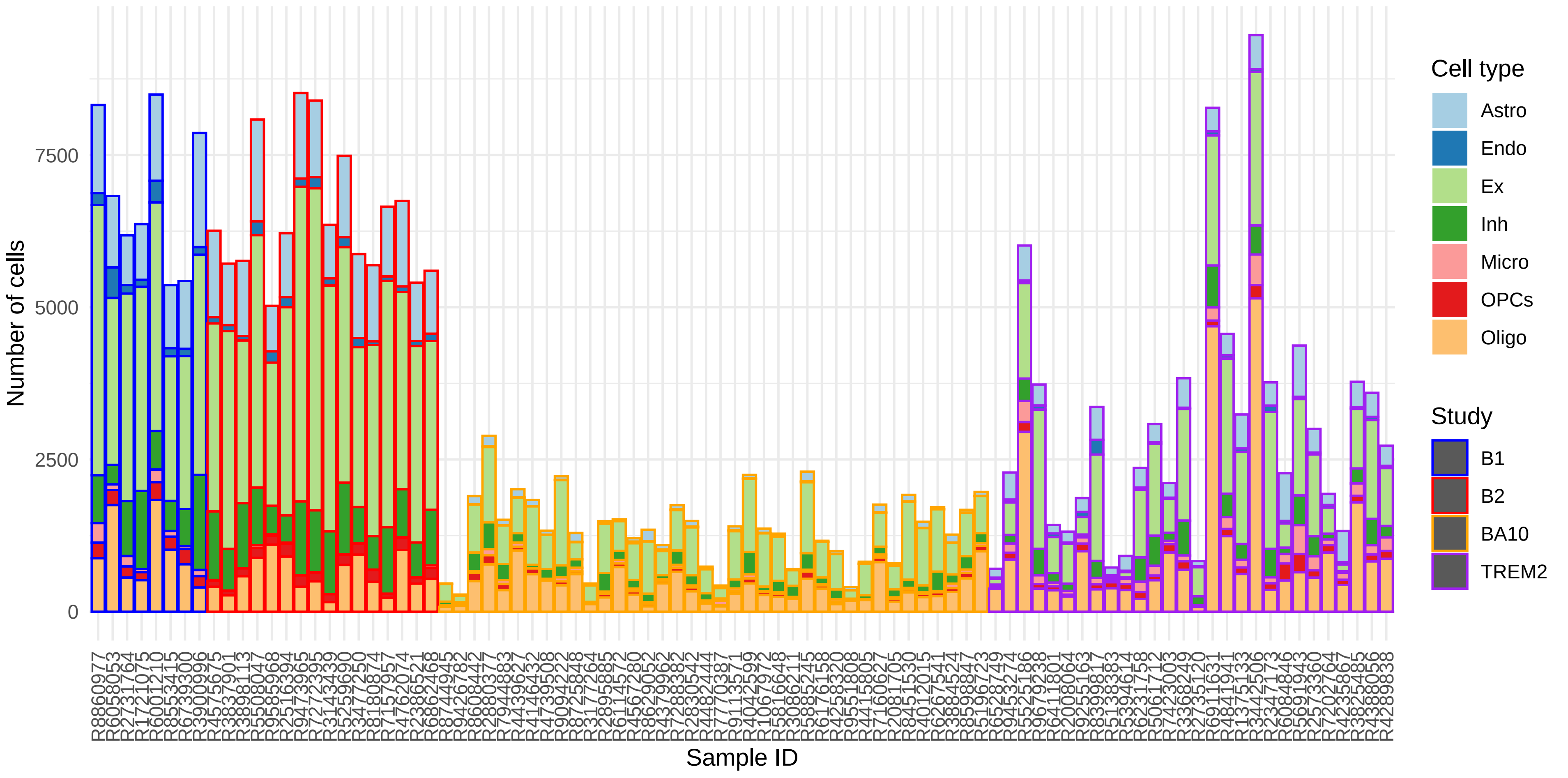

### S3 Fig

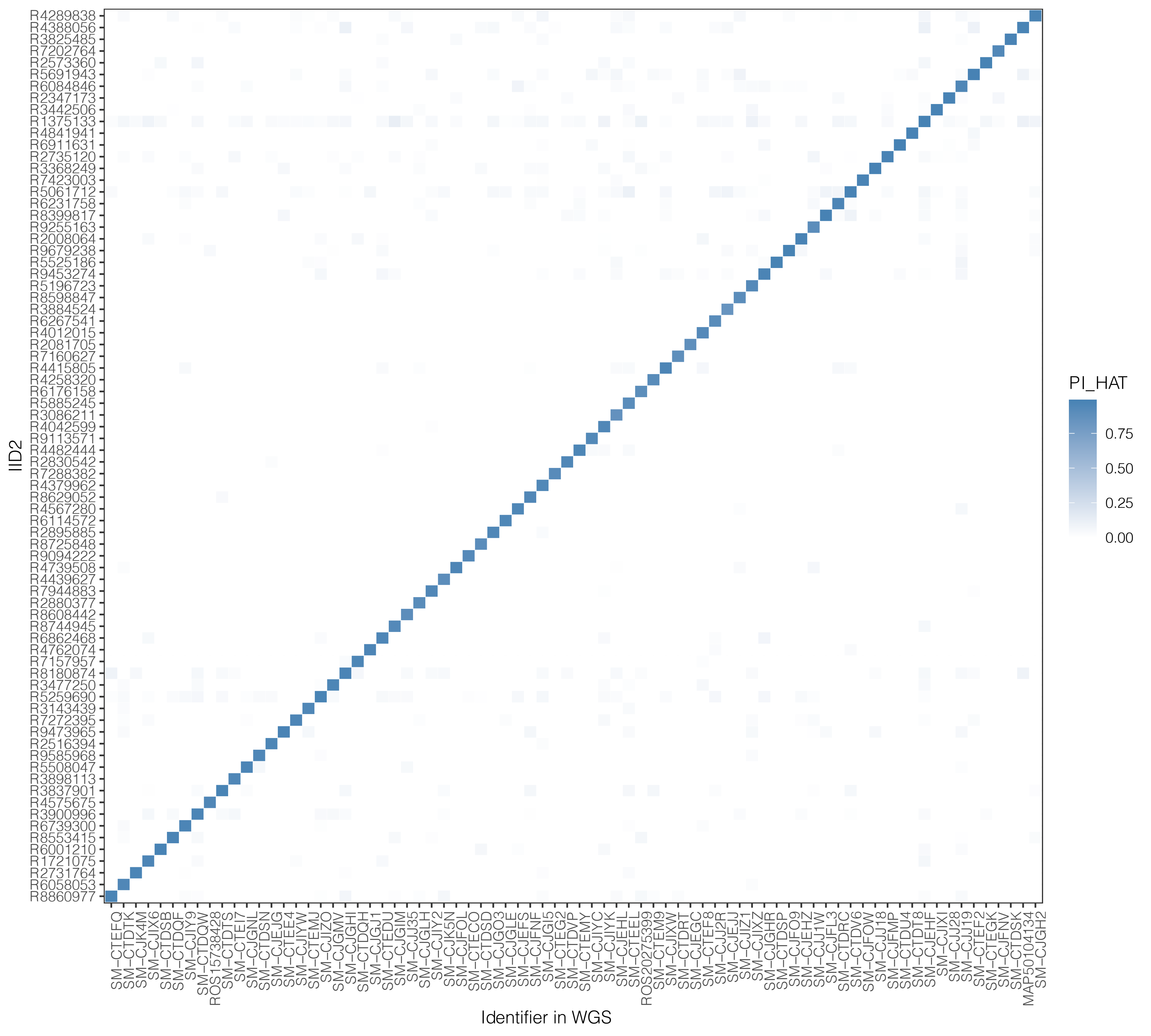

### S4 Fig

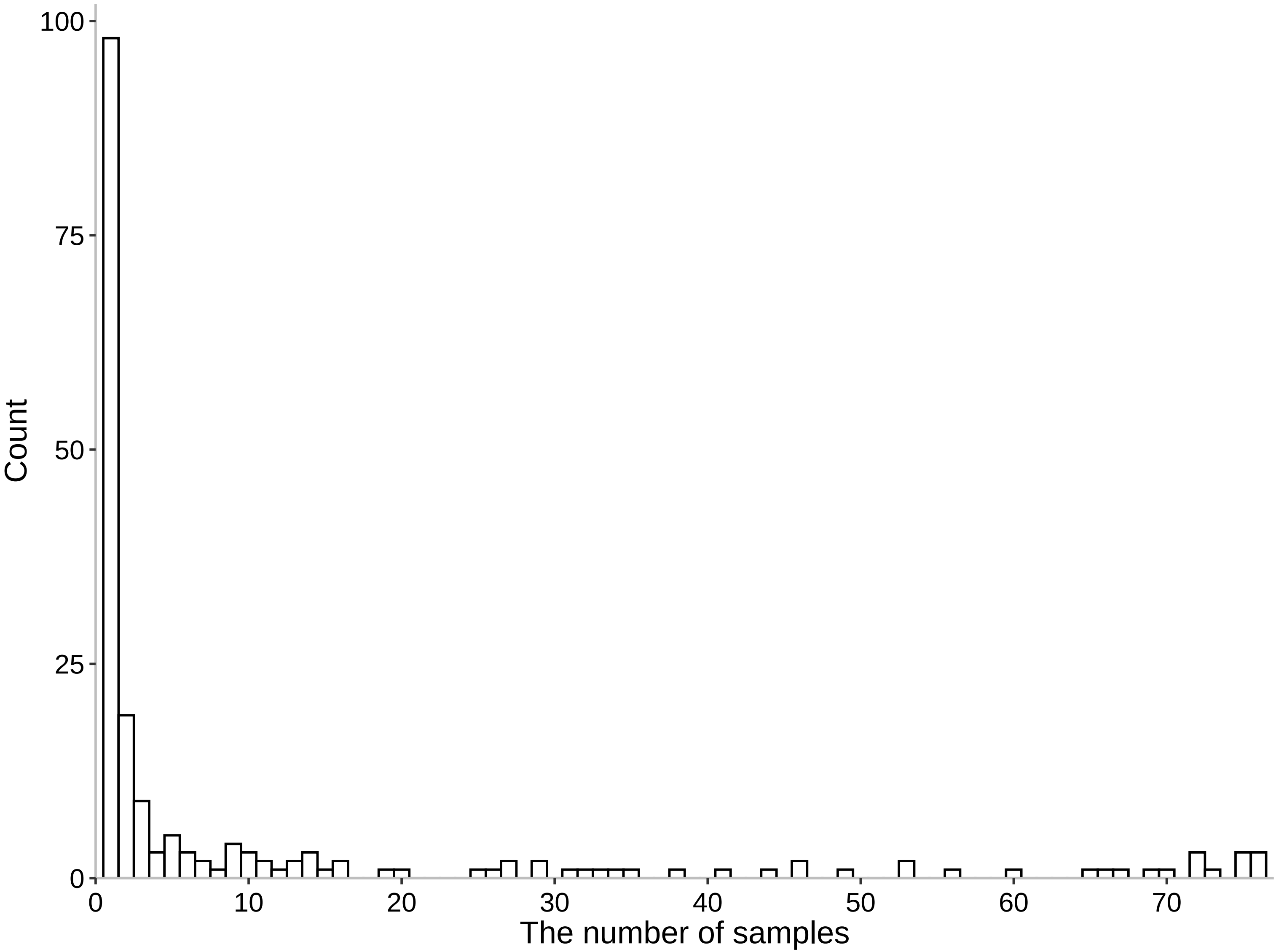

### S5 Fig

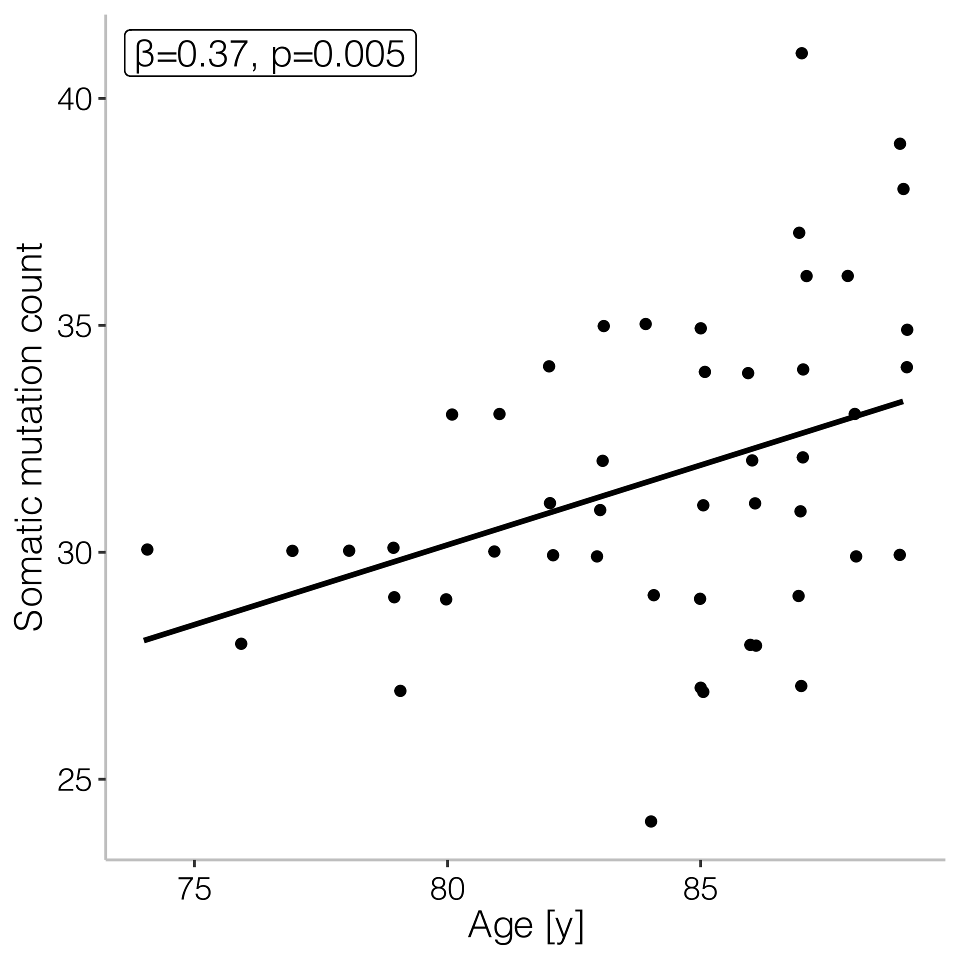

### S6 Fig

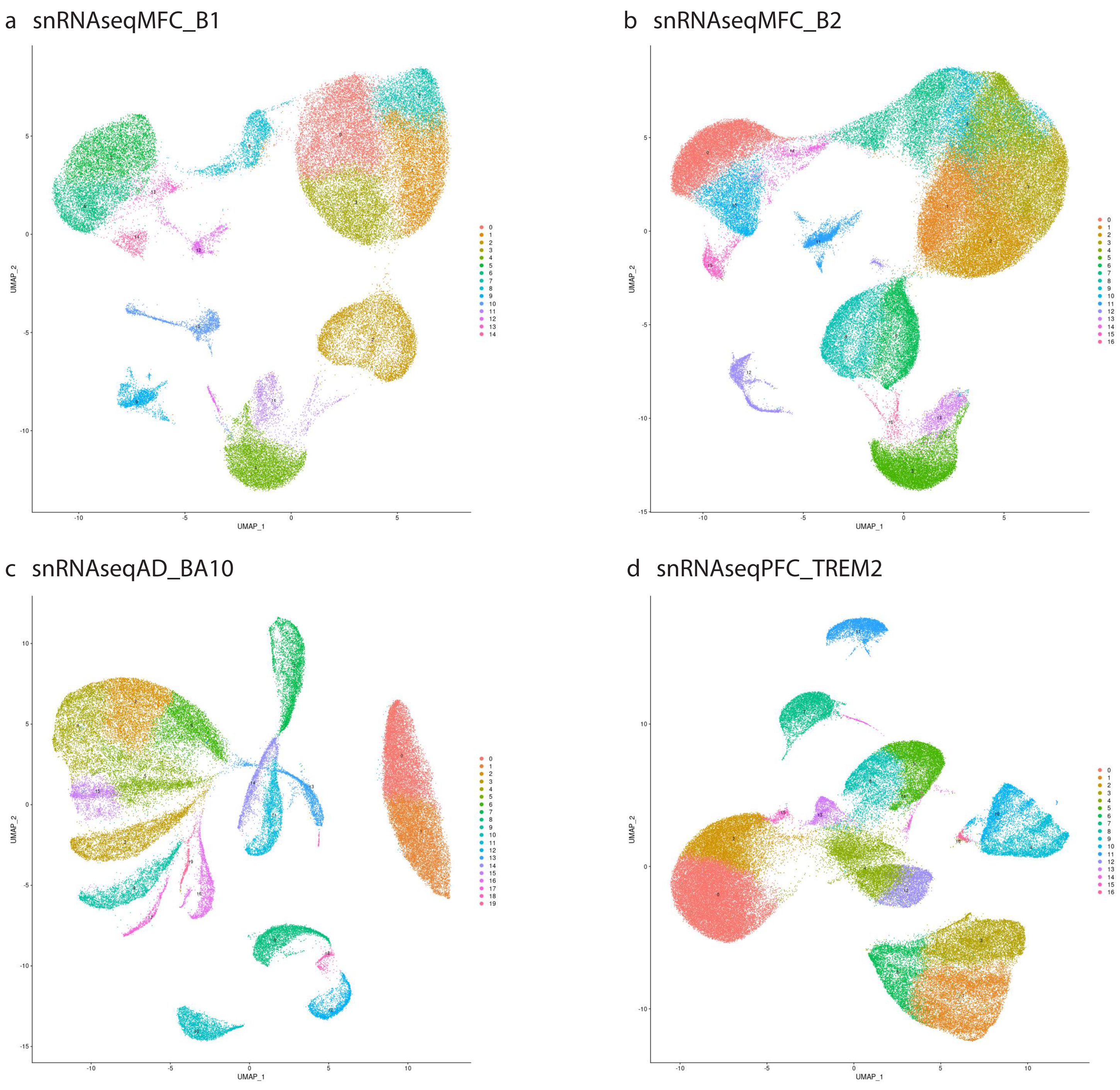

### S7 Fig

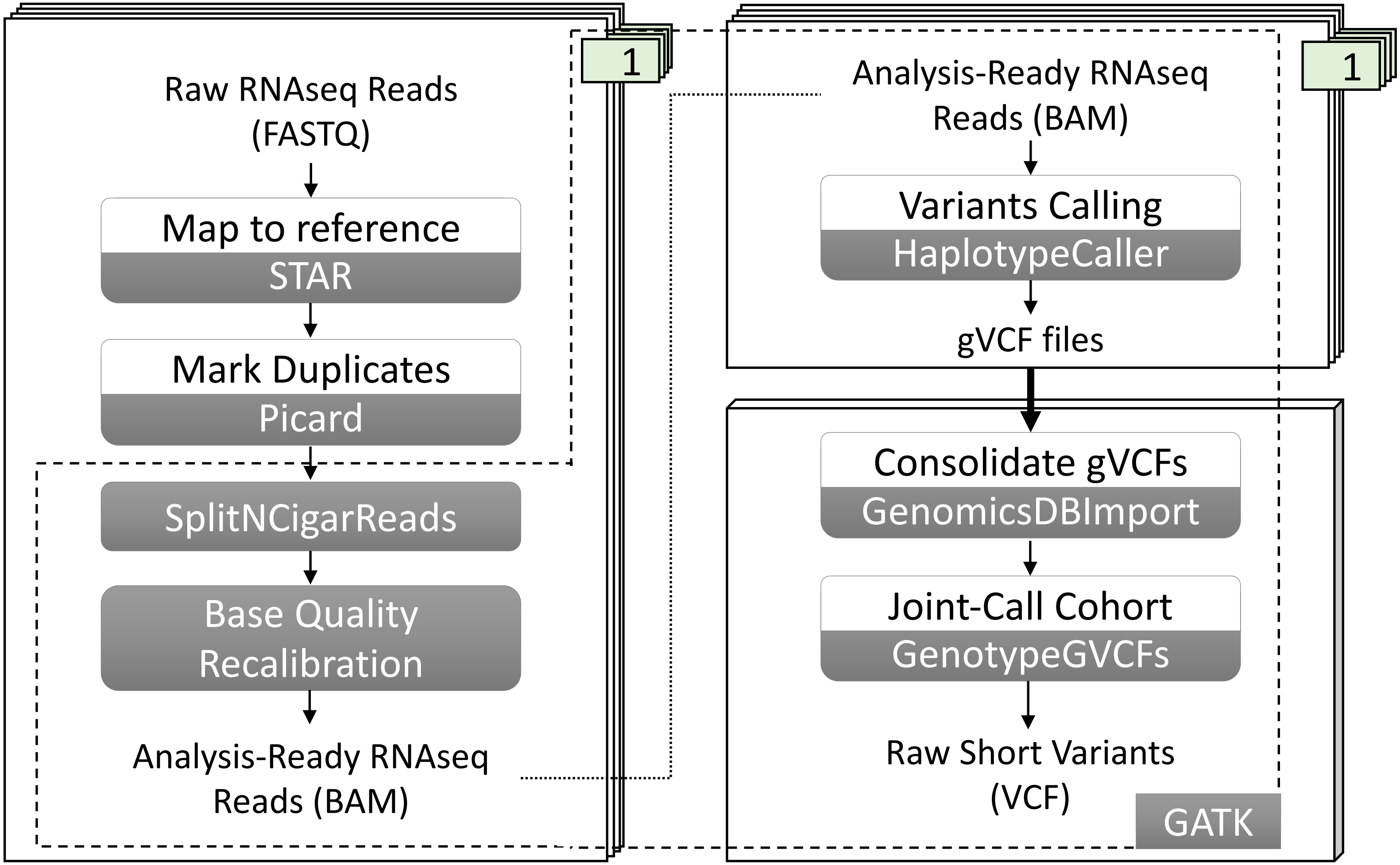
